# Interactions in abnormal synergies between the upper and lower extremities in various phases of stroke: A cohort study

**DOI:** 10.1101/2024.02.07.24302477

**Authors:** Dongwon Kim, Sung-Hwa Ko, Junhee Han, Young-Taek Kim, Yun-Hee Kim, Won Hyuk Chang, Yong-Il Shin

**Affiliations:** Shirley Ryan AbilityLab, Chicago, IL, USA; Department of Physical Medicine and Rehabilitation, Northwestern University, Chicago, IL; Department of Biomedical Engineering, University of Illinois at Chicago, Chicago, IL; Department of Rehabilitation Medicine, Pusan National University School of Medicine; Department of Rehabilitation Medicine, Pusan National University Yangsan Hospital, Yangsan, Republic of Korea; Department of Statistics, Hallym University, Chuncheon-si, Republic of Korea; Department of Preventive Medicine, Chungnam National University Hospital, Daejeon, Republic of Korea; Department of Health Sciences and Technology, Department of Medical Device Management and Research, Department of Digital Health, Samsung Advanced Institute for Health Sciences & Technology (SAIHST), Sungkyunkwan University, Seoul, Republic of Korea; Department of Physical and Rehabilitation Medicine, Center for Prevention and Rehabilitation, Heart Vascular Stroke Institute, Samsung Medical Center, Sungkyunkwan University School of Medicine, Seoul, Republic of Korea

## Abstract

**Objective:** The flexion synergy and extension synergy are a representative consequence of a stroke and appear in the upper extremity and lower extremity. Since the ipsilesional corticospinal tract (CST) is the most influential neural pathway for both extremities in motor execution, damage by a stroke to this tract could lead to similar motor pathological features (e.g., abnormal synergies) in both extremities. However less attention has been paid to the inter-limb correlations in the flexion synergy and extension synergy across different recovery phases of a stroke.

**Methods:** In this study, we used results of the Fugl-Meyer assessment (FMA) to characterize those correlations in a total of 512 participants with hemiparesis post stroke from the acute phase to 1 year. The FMA provides indirect indicators of the degrees of the flexion synergy and extension synergy post stroke.

**Results:** We found that generally, strong inter-limb correlations (r>0.65 with all p-values<0.0001) between the flexion synergy and extension synergy appeared in the acute-to-subacute phase (<90 days). But correlations of lower-extremity extension synergy with upper-extremity flexion synergy and extension synergy decreased (down to r=0.38) around 360 days after stroke (p<0.05).

**Interpretation:** These results suggest that the preferential use of alternative neural pathways after damage by a stroke to the CST enhances inter-limb correlations between the flexion synergy and extension, however a recovery of the CST or/and the functional fragmentation (remodeling) of the alternative neural substrates in the chronic phase contribute to diversity in neural pathways in motor execution, eventually leading to reduced inter-limb correlations.

## INTRODUCTION

Damage by a stroke to the ipsilesional corticospinal tract (CST) can cause a significant decrease in the capability for selective muscle activation or joint individuation. The primary reason for this decrease may be the substitution of alternative neural networks for the damaged CST ^1^. In particular, the reticulospinal tract (RST), as a dominant alternative descending pathway, connects to multiple motoneuron pools across the upper extremity and activates them simultaneously when a central command descends ^2^. Also, the cortical areas from which the RST originates, including the premotor cortex and supplementary motor area, might not have gone through fragmentation for joint individuation following stroke, causing abnormal co-activation across muscles.

Abnormal co-activation across muscles, resulting in the stereotypical flexion and extension synergy, are omnipresent in people with severe-to-moderate impairment after stroke. Those pathological phenomena are observed in the lower extremity ^3^, as well as, in the upper extremity ^4^. Lower-extremity movements are accomplished through voluntary, rhythmic or/and reactive motor controls ^5,6^. Even though complicated neural mechanisms, involving the spinal cord and limbic system, as well as the cerebral cortex, contribute to lower-extremity movements ^7,8^, it is generally accepted that those pathological synergies post stroke largely originate from the damaged CST and preferential use of alternative neural networks (see review ^7^). This could be supported by evidence from studies that demonstrate the similar recovery rates (depending on the recovery of the ipsilesional CST) ^9^ and exaggerated neural coupling ^10^ between the upper and lower extremity following a stroke. However, less attention has been paid to the interactions in the flexion synergy and extension synergy between the upper and lower extremities across different recovery phases of a stroke. The integrity of alternative pathways (i.e. RST), as well as, that of the damaged CST, could vary over the course of recovery, and accordingly, the inter-limb interactions would vary.

In this study, we investigate the time course of interactions abnormal synergies between the upper and lower extremities in participants with hemiparesis post stroke from the acute phase to 1 year. We focus on asynchronous relationships in the strengths of flexion and extension synergy expressions to minimize excitatory effects, including propriospinal excitability and spinal reflex excitability, that occur in interaction between the upper and lower extremities during voluntary activation ^11,12^. We employ the results of the Fugl-Meyer assessment (FMA). The FMA is a standardized measure suitable for multi-site investigations with high interrater reliability, providing large availability that leads to a massive database to analyze. The FMA provides indirect indicators of the degrees of the flexion synergy and extension synergy post stroke ^4^. Besides, the FMA enables us to examine improvement in the ability to activate individual muscles in the upper and lower extremities, potentially estimating CST integrity. We hypothesize that there are correlations in the extents of abnormal synergies between the upper and lower extremities, since stroke damages the CST that primarily governs movements of both extremities, and the correlations between the extremities would become weaken as the CST recovers after abnormal synergy expressions are apparent.

## METHODS

We adopted a dataset from the Korean Stroke Cohort (KOSCO) study conducted from 2012 to 2017. The study was approved by the Research Ethics Committee of Pusan National University Yangsan Hospital (Institutional Review Board number: 05-2012-057), and the participating hospitals were approved by their respective ethics committees. As a part of the study, the individuals with stroke were assessed with the FMA after 7, 90, 180, and 360 days since stroke. The inclusion criteria of the KOSCO cohort study were: (a) first-ever stroke (ischemic or hemorrhagic stroke), (b) age ≥19 years, and (c) being able to understand the purpose of the study and consent to participate. The exclusion criteria were: (a) transient ischemic attack and (b) recurrent stroke. We further imposed additional inclusion criteria for the current study: (a) hemiplegia and (b) completion of all 4 FMAs. Participants who were identified to have surgery including tracheostomy were excluded from analysis.

A retrospective approach was taken; we grouped participants into two groups based on the total score of the FMA on the paretic upper extremity (UEFM) conducted around 180 days since stroke. Participants were classified as having Mild impairment (UEFM≥43, Mild group) or Severe-to-Moderate impairment (42≥UEFM>0, Se-Mo group) ^13^. It is widely accepted that individuals with mild impairment since stroke exhibit significant recovery within the first 30-60 days, while individuals with moderate or several impairment throughout the subacute phase (< 180 days) and even the first year ^14^. Features of motor functions in those two groups are largely differentiable based on UEFM around 180 days after stroke ^15^. It is hypothesized that the Mild group preferentially uses the CST while the Se-Mo group preferentially uses alternative tracts, exhibiting apparent abnormal synergies ^16,17^.

We categorized the FMA test items into ones that assess 3 types of movements (in-flexion-synergy movement, in-extension-synergy movement and out-of-synergy movement) to grossly grade the influence of the flexion and extension synergies. For the upper extremity, part of the 33 FMA test items are categorized into subgroups including movement within the flexion synergy (6 items), movement within the extension synergy (3 items), movement mixing those synergies (3 items), and movement with no or little synergies (3 items), with the shoulder, elbow and forearm, and movements with the wrist (5 items) and fingers (7 items) ^18^. The test items requiring movement mixing synergies, movement with no or little synergies, and movements with the wrist and fingers necessitate the capability for selective muscle activation that deviates from the flexion synergy or extension synergy, which requires mature CST integrity. We grossly summed the scores of those test items to estimate CST integrity. Though hand grasping could involve the participation of the RST ^1^, our preliminary analysis confirmed that those test items were achieved in individuals with mature CST integrity. Meanwhile, we grossly summed the scores of the items for movement within the flexion synergy and movement within the extension synergy to evaluate the strength of those synergy expressions, respectively.

In the same way, for the lower extremity, part of the 17 FMA test items can be categorized into subgroups including movement within the flexion synergy (3 items), movement within the extension synergy (4 items), movement mixing those synergies (2 items), and movement with no or little synergies (2 items) ^19^ (see Table 2). We assume that the test items requiring movement mixing synergies and movement with no or little synergies need the capability for selective muscle activation that deviates from the flexion synergy or extension synergy. We grossly summed the scores of those 4 test items to estimate CST integrity.

**Table 1.**

| **Movement CLASS** | **Test item** |
| --- | --- |
| **Movement in flexion synergy** | Shoulder retraction |
|  | Shoulder elevation |
|  | Shoulder abduction |
|  | Shoulder external rotation |
|  | Elbow flexion |
|  | Forearm supination |
| **Movement in extension synergy** | Shoulder adduction/internal rotation |
|  | Elbow extension |
|  | Forearm pronation |
| **Movement mixing synergies** | Hand to lumbar spine |
|  | Shoulder flex 90° |
|  | Forearm supination/pronation |
| **Movement with little synergies** | Shoulder abduction 90° |
|  | Shoulder flex 90-180° |
|  | Forearm supination/pronation |
| **Movement with the wrist** | Wrist stability (shl at 0° and elb at 90°) |
|  | Wrist flex/ext (shl at 0° and elb at 90°) |
|  | Wrist stability (shl flex/abd and elb at 0°) |
|  | Wrist flex/ext (shl flex/abd and elb at 0°) |
|  | Circumduction |
| **Movement with the hand** | Mass flexion |
|  | Mass extension |
|  | Grasp A |
|  | Grasp B |
|  | Grasp C |
|  | Grasp D |
|  | Grasp E |

A list of individual test items of the FMA for the upper extremity that evaluate the abilities to conduct various types of movements.

**Table 2.**

| **Movement CLASS** | **Test item** |
| --- | --- |
| **Movement in flexion synergy** | Hip flexion |
|  | Knee flexion |
|  | Ankle dorsiflexion |
| **Movement in extension synergy** | Hip extension |
|  | Hip adduction |
|  | Knee extension |
|  | Ankle plantarflexion |
| **Movement mixing synergies** | Knee flexion from actively or passively extended knee |
|  | Ankle dorsiflexion (sitting) |
| **Movement with little synergies** | Knee flexion to 90°, hip at 0°,<br>balance support is allowed |
|  | Ankle dorsiflexion (standing) |

A list of individual test items of the FMA for the lower extremity that evaluate the abilities to conduct various types of movements.

We did not count on the reflex test items, because we found that scoring in those items was not correlated with the degree of impairment. The other test items were excluded from analysis which were found to not be correlated with the test items addressed above ^20^.

To estimate the relative proportion in scoring in test items between out-of-synergy movements and in-synergy movements, we calculated Theta, defined as the angle of a right triangle formed by two legs (Fig. 1): the summed score of in-synergy test items normalized to the possible maximum score and the summed score of out-of-synergy test items normalized to the possible maximum score. Thetas were calculated using the inverse trigonometric function of the tangent function. Thetas were calculated for the flexion synergy and extension synergy, respectively. The value of Theta approaching 0 degrees indicates scoring in in-synergy test items is much higher than that in out-of-synergy test items, while the value of Theta approaching 90 degrees indicates scoring in out-of-synergy test items is much higher than that in in-synergy test items. The value of Theta approaching 45 degrees indicates balanced scoring between the two types of test items.

**Figure 1.**
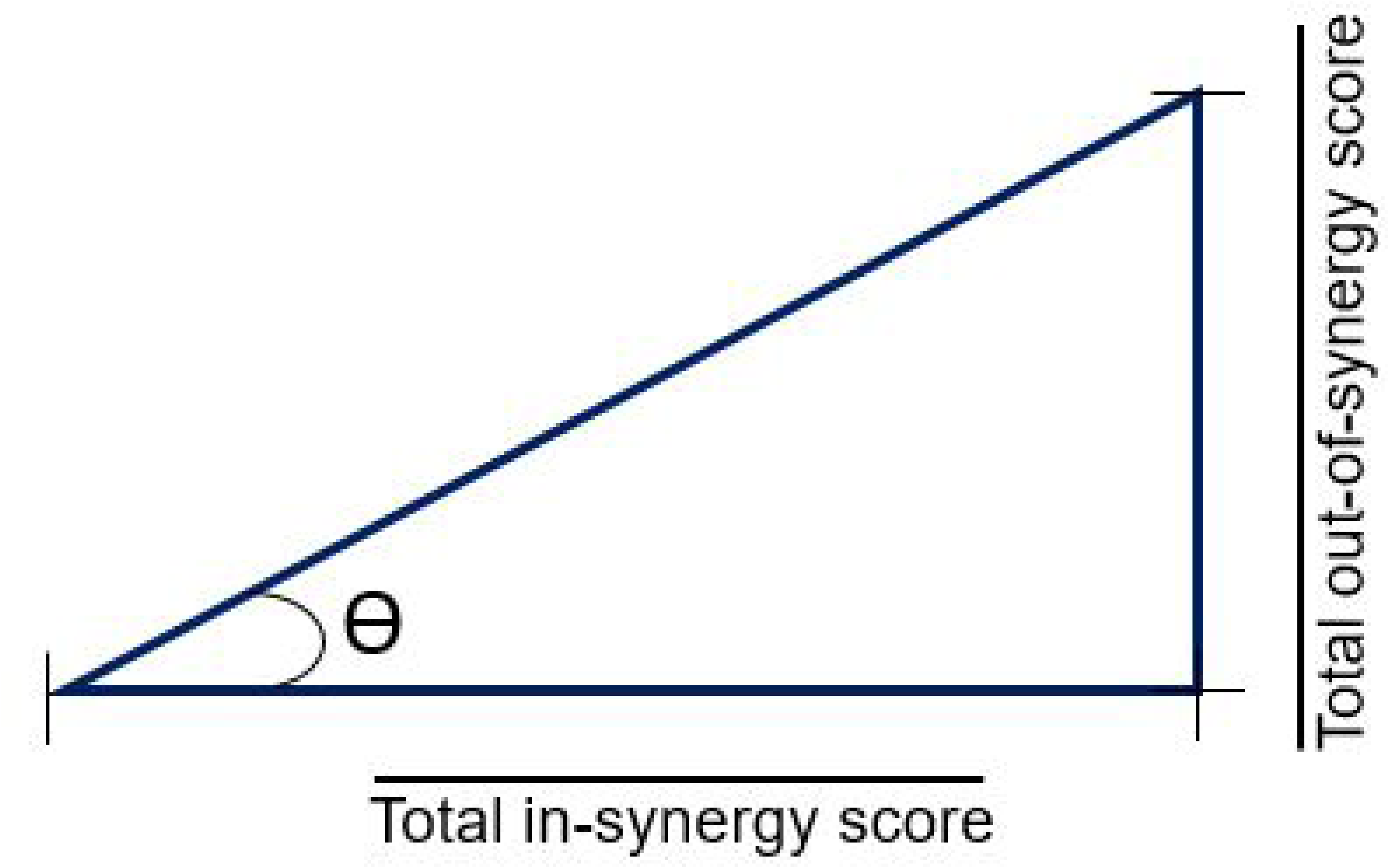
Definition of Theta (Ɵ).

We investigated longitudinal changes in FMA scores using repeated-measures ANOVA. When the sphericity assumption was violated, the Greenhouse–Geisser method was used to adjust p-values. A parametric correlation analysis (Pearson’s correlation) was employed to investigate the trends of changes in the summed score of out-of-synergy test items between the upper extremity and lower extremity. We used this analysis to investigate correlations in Theta between the extremities. Correlation coefficients were compared with the Bonferroni correction. Statistical analyses were performed with SPSS (Windows v.18, SPSS Inc.). The significance level was set at 0.05.

## RESULTS

A total of 512 individuals completed assessments around 7, 90, 180, and 360 days after stroke. Ninety five individuals were identified to be in the Se-Mo group and 417 individuals in the Mild group. There were no significant differences in age, sex (ratio of males versus female) and time since stroke at each assessment between the two groups. Participant demographics are presented in Table 3.

**Table 3.**

|  | Mild group | Se-Mo group |
| --- | --- | --- |
| N | 417 | 95 |
| Age | 72.0±12.9(s.d.) years | 76.1±13.4(s.d.) years |
| Sex | 277 Male /140 Female | 54 Male/41 Female |
| First-ever | 417 Yes | 95 Yes |
| Stroke type | 363 Ischemic/<br>54 Hemorrhage | 66 Ischemic/<br>29 Hemorrhage |
| Time since stroke |  |  |
| 1<sup>st</sup> assessment | 7.1±4.9(s.d.) days | 6.9±2.3(s.d.) days |
| 2<sup>nd</sup> assessment | 90.2±5.6(s.d.) days | 92.4±8.3(s.d.) days |
| 3<sup>rd</sup> assessment | 178.8±7.8(s.d.) days | 180.4±9.2(s.d.) days |
| 4<sup>th</sup> assessment | 360.7±7.5(s.d.) days | 360.5±5.0(s.d.) days |

Participant demographics (s.d. stands for standard deviation).

The Mild group showed improvements in scoring for the flexion synergy, extension synergy and out-of-synergy, in both extremities from 7 days to 90 days following stroke (p<0.05), as shown in Fig. 2. Meanwhile the Se-Mo group showed the same trend of improvement during that period (p<0.05), except for the out-of-synergy test items in the upper extremity. We observed significant motor improvements in scoring for the flexion synergy, extension synergy and out-of-synergy, in both extremities in the Se-Mo group, after 180 days.

**Figure 2.**
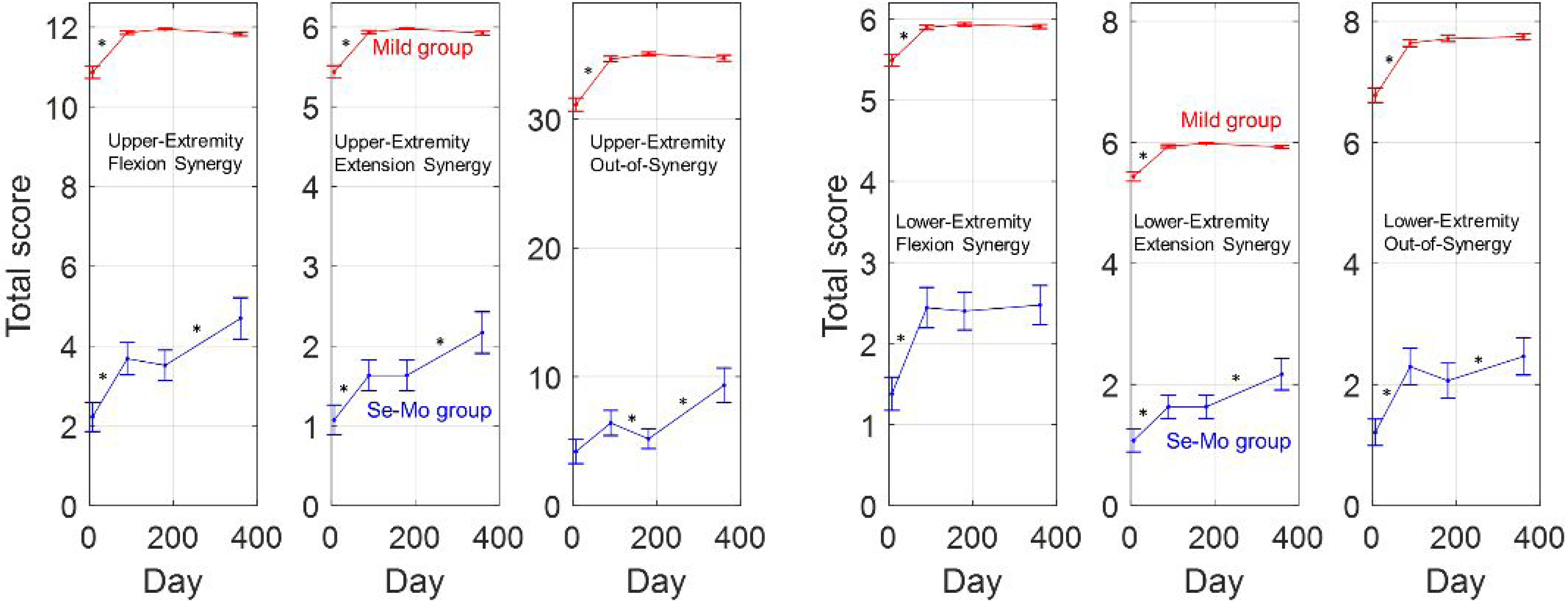
Time evolutions of the total score of in-synergy (flexion and extension synergies) test items and total score of out-of-synergy test items in the Mild group and Se-Mo group. Error bars are standard error and asterisks indicate a significant difference in mean between two assessments (p<0.05).

We found significant correlations (p<0.05) in the normalized increment in the total score of the out-of-synergy test items of the lower extremity versus the upper extremity between time points of assessment (Fig. 3). We note that the recovery rate of the upper-extremity capability for the type of out-of-synergy movement is greater than that of the lower-extremity capability after 180 days to 360 days (regression line slope=0.49), as compared to the other periods (p<0.05).

**Figure 3.**
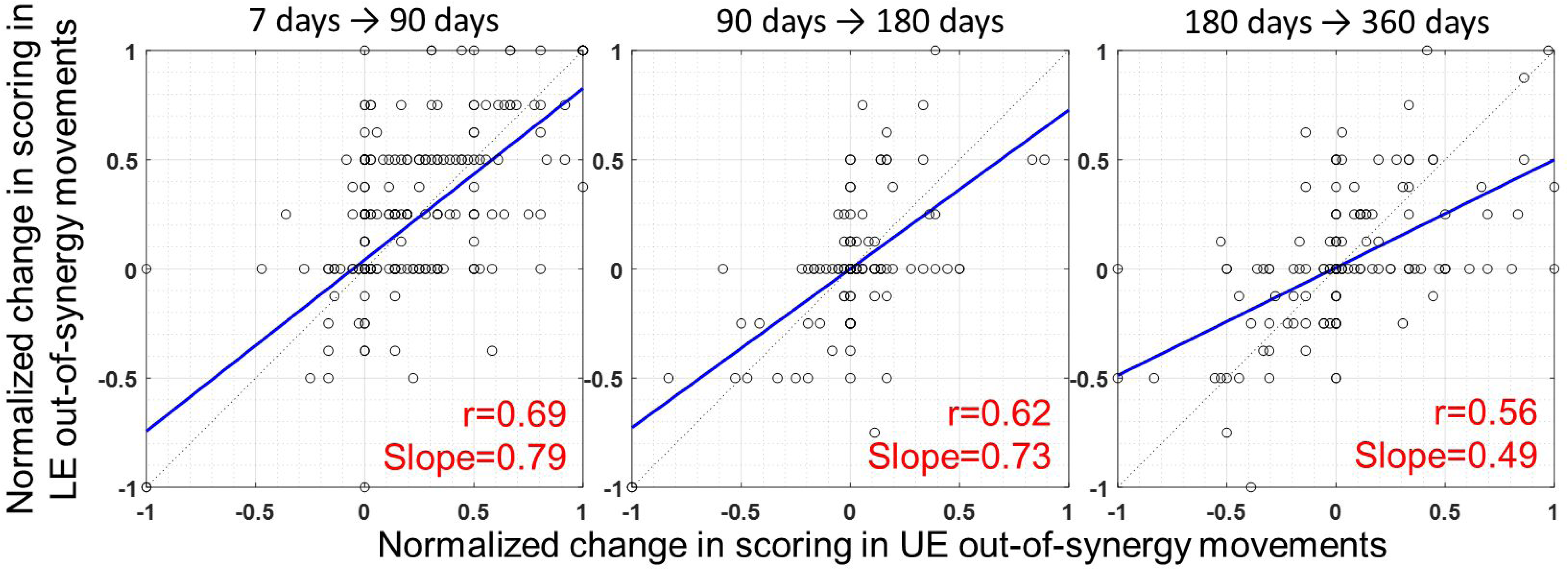
Distributions of the increment in the total score of the out-of-synergy test items for the lower extremity normalized to its possible maximum score versus the increment in the total score of the out-of-synergy test items for the upper extremity normalized to its possible maximum score between time points of assessment. The blue solid lines are the regression lines and dotted lines are lines with a slope of 1.

In the Mild group, Thetas approached 45 degrees for the upper extremity and lower extremity throughout all phases of stroke, suggesting their balanced scoring between the in-synergy (flexion or extension synergies) test items and total score of out-of-synergy test items (Fig. 4). The Se-Mo group reached values below 25 degrees about all Thetas in the acute phase. These results imply that the scoring distribution in this group was substantially biased towards in-synergy test items. After the acute phase, the values of all Thetas significantly increased, mainly during the period from 7 days to 90 days (p<0.05), but those did not reach the levels of the Mild group (p<0.05). This suggests that the scoring distribution in the Se-Mo group was still biased towards in-synergy test items in all phases of stroke we considered.

**Figure 4.**
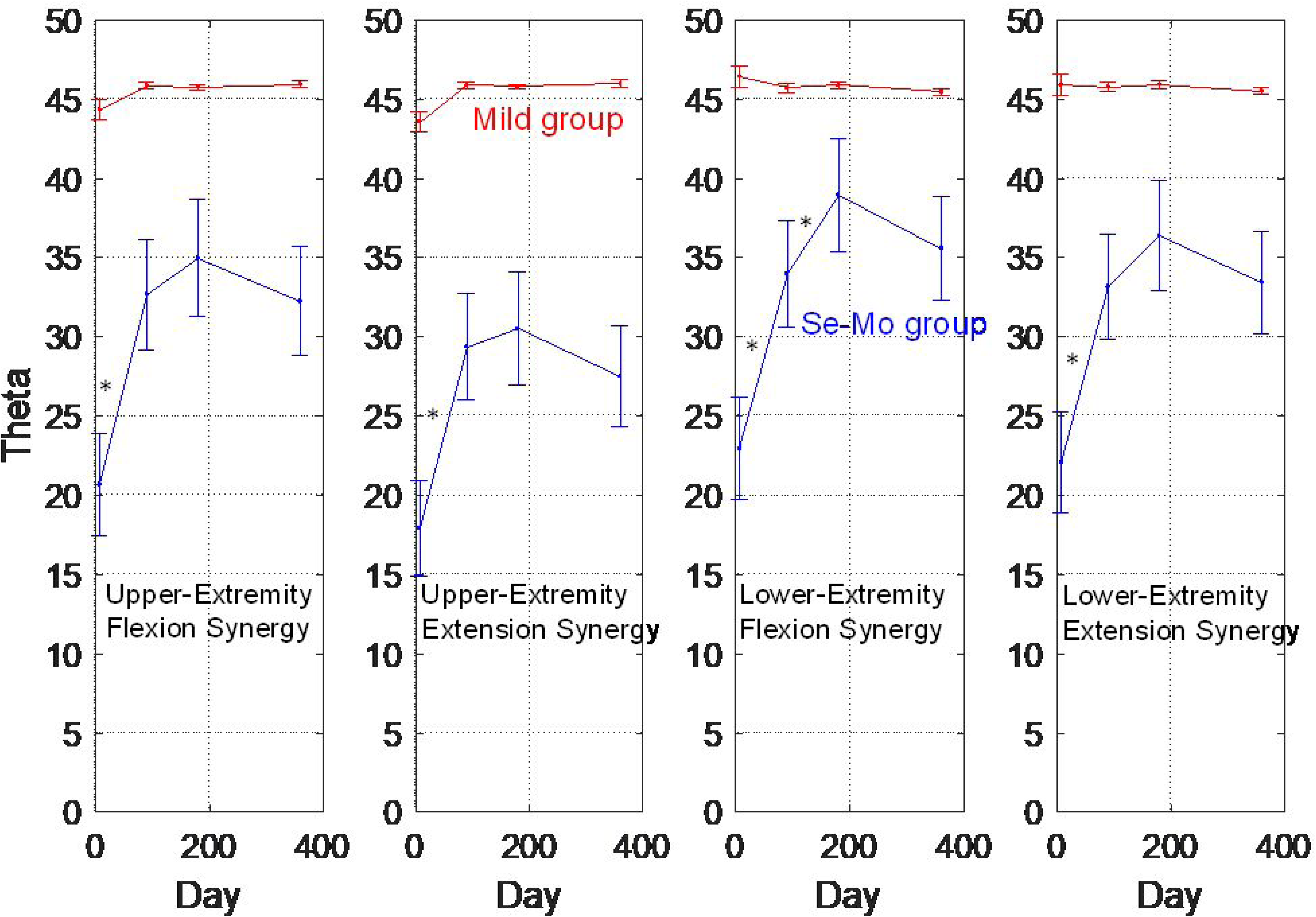
Time evolutions of Thetas for the upper extremity and lower extremity in the Mild group and Se-Mo group. The value of Theta approaching 0 degrees indicates scoring in in-synergy test items is much higher than that in out-of-synergy test items, while the value of Theta approaching 45 degrees indicates balanced scoring between the two types of test items. Error bars are standard error and asterisks indicate a significant difference in mean between two assessments (p<0.05).

As shown in Fig. 5, we observed strong inter-limb correlations (r>0.65 with all p-values<0.0001) between the flexion synergy and extension synergy around 7 days and 90 days after stroke. Meanwhile we found that correlations of lower-extremity extension synergy with upper-extremity flexion synergy (r=0.48) and extension synergy (r=0.38) significantly decreased around 360 days after stroke (p<0.05).

**Figure 5.**
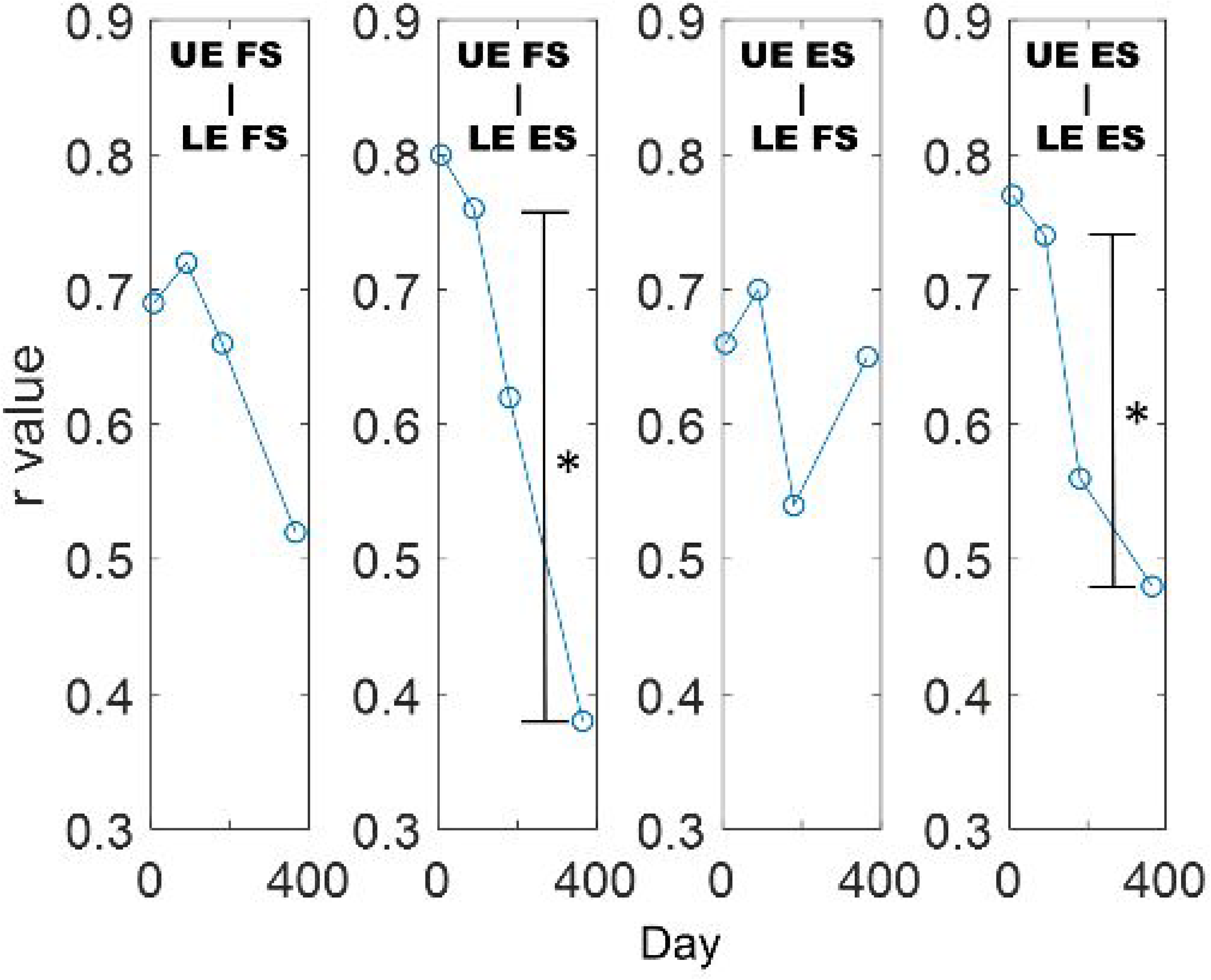
Time evolutions of correlation coefficients between Thetas for the upper extremity (UE) and lower extremity (LE) (flexion synergy (FS) and extension synergy (ES), respectively) in the Se-Mo group. Asterisks indicate a significant difference (p<0.05/6: Bonferroni correction (6 is the number of comparisons across 4 phases)).

## DISCUSSION

This study addressed 1) the time evolutions of recovery of motor function in the upper and lower extremities following stroke, differentiating individuals based on the degree of impairment assessed around 180 days after stroke, 2) the asynchronous relationships in the strengths of flexion and extension synergy expressions across various phases of stroke, and 3) the correlation in improvement in the ability to activate individual muscles between the upper and lower extremities. In particular, investigating asynchronous relationships can rule out excitatory effects, including propriospinal excitability and spinal reflex excitability, that occur in interaction between the upper and lower extremities during voluntary activation ^11,12^.

**Fig. 2** shows different time courses of recovery depending on the severity of impairment assessed in the chronic phase (180 days since stroke). Spontaneous recovery largely depends on CST connectivity within 2 weeks after the advent of a stroke ^15^. If the initial CST connectivity is sufficient (i.e., the Mild group), the neural system preferentially uses the ipsilesional CST, and hand and arm dexterity recovers within 30-60 days ^14,21^. For the Mild group, we observe significant increases in scoring in the out-of-synergy test items for both the upper and lower extremities between 7 and 90 days. We also see significant increases in scoring in the flexion-synergy and extension-synergy test items for the upper and lower extremities between 7 and 90 days. This indicates that the types of flexion-synergy and extension-synergy movements are highly mediated via the CST in this group. Thetas remain around 45° across all phases of stroke (**Fig. 4**), suggesting that improvements in the abilities to conduct all types of movements are in-phase, which is perhaps because all types of movements are mediated via the CST. If the initial CST connectivity is not sufficient enough to convey motor commands (i.e., the Se-Mo group), alternative pathways are hypothesized to convey motor commands from the cortices. Motor recovery with strong reliance on alternative tracts typically continues over the first year of stroke, implying that it takes more time for more impaired persons to adapt to use of compensatory mechanisms for the damaged CST and/or achieve functional fractionation of the alternative neural pathways ^14,21,22^. Shifts in inter-hemispheric lateralization may occur towards the contralesional hemisphere and changes in representational maps may occur around the infarcted zone ^23^. The results of Thetas for the Se-Mo group that remain far below 45° across all phases (**Fig. 4**) support this; the type of in-synergy movements outperforms the type of out-of-synergy movements, suggesting the preferential use of the RST and contralesional hemisphere.

Scoring in the out-of-synergy test items for the upper extremity decreases between 90 and 180 days. This phenomenon could be explained by the functional upregulation of the RST which leads to enhanced abnormal synergies appears in the chronic phase ^24,25^. RST upregulation could reduce the ability to conduct the type of out-of-synergy movements for a while, possibly leading to a significant decrease in scoring in the out-of-synergy test items. It is not clear which neural substrate primarily leads to increases in scoring in the out-of-synergy test items between 180 and 360 days. We can raise the possibility of functional fragmentation (remodeling) of the alternative neural substrates (i.e. contralesional cortices and RST). The same trends are observed in scoring in the flexion-synergy movement test items and in scoring in the extension-synergy test items. Several studies demonstrated that structural reorganization of the contralesional cortices undergoes following stroke and contributes to motor improvement, possibly promoting joint individuation ^26,27^. Also, it is possible that the recovery in the CST contributes to those increases in scoring. Our modeling study demonstrated that corticospinal networks become optimized slowly while reticulospinal networks become optimized with priority (unpublished). Once optimization of reticulospinal networks completes, the neural system optimizes the remaining available networks (i.e. corticospinal networks).

Overall, our results suggest that the same neural substrate (i.e. CST or RST) may work for both extremities ^7,10^. We observe similar trends of recovery in the upper and lower extremities in the Se-Mo group as well as the Mild group, which is generally in agreement with the findings of studies ^9,28^. The increases in scoring in the out-of-synergy test items for the upper extremity are significantly correlated with those for the lower extremity across phases (p<0.05, **Fig. 3**). Changes in scoring in the in-synergy test items are also in-phase between two extremities (**Fig. 2**).

The highlight of the study is the interactions in the flexion synergy and extension synergy between the upper and lower extremities across different recovery phases of a stroke. Only a limited number of studies addressed synchronous interactions in muscle activation between the upper and lower extremities post stroke ^10,29,30^. Studies found that involuntary flexion in the upper extremity occurs while walking ^10,29,30^. Also, involuntary activation of an extensor (i.e. the rectus femoris) in the lower extremity occurs while finger flexing ^10^. Though we focused on asynchronous interactions, our results are in agreement with those of those studies. In particular, our results show significant correlations between the upper-extremity flexion synergy and lower-extremity extension synergy (p<0.05, **Fig. 5**). Though excitability of the vestibulospinal networks might be one of the reasons for such interactions, the prosperity of the RST following stroke could be the primary reason ^2,10^. As mentioned in a study ^10^, an effort to counteract the gravitational loading requires the RST to be excited for the lower extremity as well as the upper one in individuals with severe-to-moderate impairment ^1,2,31^. The flexion synergy in the upper extremity and the extension synergy in the lower extremity are highly correlated, both of which are excited when counteracting the gravitational loading. However, increased reliance on the RST following stroke also enhances other synergies that are irrelevant to the gravitational loading. We observe other significant correlations of the flexion synergy and extension synergy of the upper extremity with those of the lower extremity (p<0.05, **Fig. 5**). Studies demonstrated that the contralesional RST enhances the flexion synergy ^32^ and the ipsilesional RST does the extension synergy in the case of the upper extremity ^33^. It could be that those two pathways work for the flexion synergy and extension synergy in the lower extremity, respectively, and that those interact. Indeed, knee extension causes involuntary activation of extensors in the upper extremity ^10^. Increased integrity and use of the reticulospinal networks may lead to high correlations in the flexion synergy and extension synergy between the upper and lower extremities.

Note that we observe significant decreases in the correlations between the upper-extremity flexion synergy and lower-extremity extension synergy and between the upper-extremity extension synergy and lower-extremity extension synergy, from 90 days to 360 days after stroke (p<0.05, **Fig. 5**). These results may originate from functional fragmentation of the RSTs and contralesional cortices on executing movements with the upper and lower extremities ^26,27^. Or the recovery of the CST may contribute to those significant decreases in the correlations. Those two factors could lead to diversity in neural pathways in motor execution, eventually leading to reduced inter-limb correlations. Indeed, the ability to activate individual muscles in individuals with severe-to-moderate impairment is promoted after 180 days (**Fig. 2**).

This study exposes several limitations. The first limitation originates from the nature of the FMA. The FMA simply relies on the rater’s knowledge and experience, and accordingly ratings could be subjective, even though a guideline for consistent instructions was given. However, studies showed high inter-rater reliability for both extremities ^34–37^. Also we believe that our large amount of data mitigates the influence of inter-rater variability. The second limitation is the relatively less number of test items of the FMA for the extension synergy in the upper extremity and the flexion synergy in the lower extremity in comparison to that for the upper extremity. The evaluation on the abilities to conduct those types of movements is coarser; those are assessed with only 3 items. However, we believe that the abilities to conduct those types of movements can be graded sufficiently differentially based on ratings (0-6 points). The third limitation is as to whether our data capture spontaneous recovery alone, not affected by therapeutic interventions. Motor recovery probably occurs through a combination of spontaneous biological processes and activity- or use-dependent processes ^38^. We did not control for physical therapies individuals received. However, the separation of spontaneous biological processes from the activity- or use-dependent processes in the subacute phase have not been fully understood ^39^. Motor function undergoes fairly predictable phases of recovery over the first 6 months after stroke, regardless of types of therapeutic intervention ^40^.

## Acknowledgement

This work was supported by the Research Program funded by Korea Disease Control and Prevention Agency (3300-3334-300-260-00, 2013-E33017-00, 2013E-33017-01, 2013E-33017-02, 2016-E33003-00, 2016-E33003-01, 2016-E33003-02, 2019-E3202-00, 2019-E3202-01, 2019-E3202-02, 2022-11-006). Kim was supported by NIH NINDS (3R01NS053606).We express our gratitude to Dr. Margit A. Murphy at Gothenburg University for her insights.

## Data availability

Data will be available on reasonable request.

## Contributions for each author

Dongwon Kim

Conceptualization, Data curation, Formal analysis, Investigation, Methodology, Software, Supervision, Validation, Visualization, Writing – original draft, Writing – review & editing

Sung-Hwa Ko

Funding acquisition, Project administration, Writing – review & editing

Junhee Han

Funding acquisition, Project administration, Writing – review & editing

Young-Taek Kim

Funding acquisition, Project administration, Resources, Writing – review & editing

Yun-Hee Kim

Funding acquisition, Project administration, Resources, Writing – review & editing

Won Hyuk Chang

Funding acquisition, Project administration, Resources, Writing – review & editing

Yong-Il Shin

Funding acquisition, Project administration, Resources, Writing – review & editing

